# A Comparative Analysis of US and UK Site Self-Assessment of Practices in Recruiting and Meeting the Needs of Diverse Patient Populations in Clinical Trials

**DOI:** 10.1101/2023.11.03.23297984

**Authors:** Peter Phiri, Diana Foster, Bupendra Shah, Sana Sajid, Lynis Lewis, Shanaya Rathod, Gayathri Delanerolle, Jian Qing Shi

## Abstract

**Background:** The comparative analysis of site practices in recruiting and outreach of diverse patient populations in clinical trials is needed for a harmonious approach to navigating solutions for improving patient diversity globally. With the availability of the Diverse Site Assessment Tool (DSAT), such analyses have become feasible.

**Methods:** This study presents a comparison of self-reported data related to diversity best practices using the DSAT by members of clinical trials research sites from three distinct cohorts in the United States (US) and United Kingdom (UK). The Diverse Site Assessment Tool (DSAT) is a 25-item self-report measure, exploring best practices for the recruitment of diverse patient populations during clinical trials. The DSAT was administered online via the Qualtrics XM platform. DSAT data for each of these cohorts were retrieved and scored and individual item means were used to conduct reliability, descriptive and inferential statistical analysis.

**Findings:** Results indicate that the DSAT is an exceptionally reliable instrument for self-assessment of diversity best practices. Results also showed that among the three DSAT sections, the section of patient focused services shows the lowest scores regardless of the cohort and that DSAT section scores were considerably different between the 2 US cohorts as well as between the US and UK cohorts. Overall, DSAT scores were higher for US cohorts than for the UK cohort.

**Interpretation:** Different stakeholders from the US and UK might want to examine the findings of this study carefully and discuss its implications for their own country, sites within it and discuss how harmonized efforts for diversity in clinical trials can be established.

**Funding:** Funded by the The Society of Clinical Research Sites, USA.

## Background

Research on clinical trials has well established that the demographics of participants in clinical trials are not necessarily reflective of the actual population who will be the end users of the outcome product following decades of intensive research. Regulatory agencies such as the Food and Drug Administration has a long-documented history of trying to expand the nature of participants in clinical trials^1^. Yet, challenges related to diversity of participants in clinical trials remain and with efforts towards harmonisation, regulatory agencies such as the Food and Drug Administration (FDA), health systems agencies such as the National Health Service have further stepped up to address the inequities of representation. Frameworks, guidance, incentives, white papers, programs, and materials^2-14^ have either been developed or are being developed to help improve diversity of participants in clinical trials.

One of the positive forces towards diversity in clinical trials and a notable resource is the Diversity Site Assessment Tool (DSAT) developed by Dr. Foster and partners associated with the Diversity Awareness Program at the Society for Clinical Research Sites, an organisation representative of the needs of clinical research sites globally ^15,16^. The DSAT is a 25-item self-assessment tool based on identifying best practices related to recruitment of diverse patient populations during clinical trials and is considered to be an industry gold standard that can be completed by members of a clinical trial site to identify the capacity of their site for recruiting and meeting the needs of diverse patients in clinical trials conducted by their site. The 25 items are divided into three sections: a 10-item Site Overview section, a 9-item Site Recruitment and Outreach section and a 6-item Patient Focused Services section. Each of these sections are scored on a 6-point scale (1-Hardly ever; 6-Always), allowing site members to gain an comprehensive self-evaluation of the extent to which they are engaged in best practices associated with diversity in clinical trials. The scoring mechanism of the DSAT also provides an opportunity for a comparative analysis among and across sites, locally as well as globally. Given established reliability and validity of the DSAT, it has been argued that it provides the global clinical trials industry a tremendous opportunity to use a continuous quality improvement and systems-based approach to address recruitment of diverse populations in clinical trials^15-17^. Since its inception, the DSAT has been discussed at numerous forums for widespread adoption so that the clinical trials industry can benefit from its findings and tailored resources can be directed to help improve diversity in clinical trials.

Research using DSAT is at a nascent stage as only some analysis of data from US clinical trials site has been published. More robust studies especially those that provide insights into the current practices at clinical sites in other countries and their needs pertaining to improving diversity of participants in the clinical trials are much needed. With increased globalization of pharmaceutical product development and testing and the needs associated with harmonization, studies comparing clinical trial site’s best practices in diversity recruitment become necessary and important for a global dialogue and effort towards improving diversity in clinical trials. This study aims to address this gap in the literature by comparing DSAT data completed by members of clinical trials research sites from three cohorts, two from United States and one from United Kingdom.

## Methods

### Study Design

A cross-sectional observational study was delivered digitally. Quantitative data was collected through an online survey questionnaire using the Qualtrics Core XM platform to better understand the understand knowledge, expertise, and best practices to meet the needs of the diverse population in the UK. This study is based on a retrospective cohort analysis of DSAT data completed by three cohorts of clinical trials site members: two from the USA and one from the UK. The survey link for DSAT was distributed over a 1-year period to research active sites. US cohort 1 were provided access to the DSAT starting April 2021; US cohort 2 were provided access starting March 2021; UK cohort were provided access starting September 2022. Data was extracted in March 2023 for each of the cohorts, results were deidentified and imported to MS Excel to allow for comparison of DSAT section scores and individual item means. JASP, an open-source statistics program, was utilized to conduct reliability, descriptive and inferential statistical analysis^18^.

### Materials

The survey consisted of a section collecting background information about the participant and their site, followed by the Diversity Site Assessment Tool (DSAT). The DSAT is a 25-item self-report measure consisting of three sections which are indicators of best practices for the recruitment of diverse patient populations during clinical trials: Site Overview, Site Recruitment and Outreach, and Patient Focused Services. Each of the total 25 items on DSAT would require a site representative participant to self-report on each item on a 6-point scale. Each item from the DSAT is scored based on 1 point for answering “Hardly ever (<or =5% of the time)”, 2 points for answering “Rarely (6-24% of the time), 3 points for answering “Sometimes (25-49% of the time), 4 points for “Often (50-74% of the time), 5 points for “Nearly Always (75-94% of the time) and 6 point for answering “Always (95% or more of the time)” to each of the diversity best practice specific questions. The total DSAT scale scores have potential to range from 0-150 with higher scores indicating greater use of diversity best practices. The questionnaire was administered and completed online using Qualtrics XM platform in the following format:

SECTION 1: Representative participant and site demographic information.

SECTION 2: Site Overview (10-items)

SECTION 3: Site Recruitment and Outreach (9-items)

SECTION 4: Patient Focused Services (6-items)

The survey took approximately 15 minutes to complete with participants able to omit questions they did not wish to complete.

### Participants

Participants for the first US cohort consisted of members from the non-profit, global representative of clinical sites. Participants for the second US cohort consisted of members associated with an independent, full service clinical trials diversity organization. Participants for the cohort from the United Kingdom were members of NHS and non-NHS sites throughout UK who were approached through multifaceted social media platforms, NIHR networks and other research networks and registers.

## Results

Data for a total of 853 participants was received, with a significantly larger participation from the USA. A total of 224 participants took part in the study. Table 1a and 1b outline the demographic characteristics of the UK and US samples, respectively.

**Table 1a.**
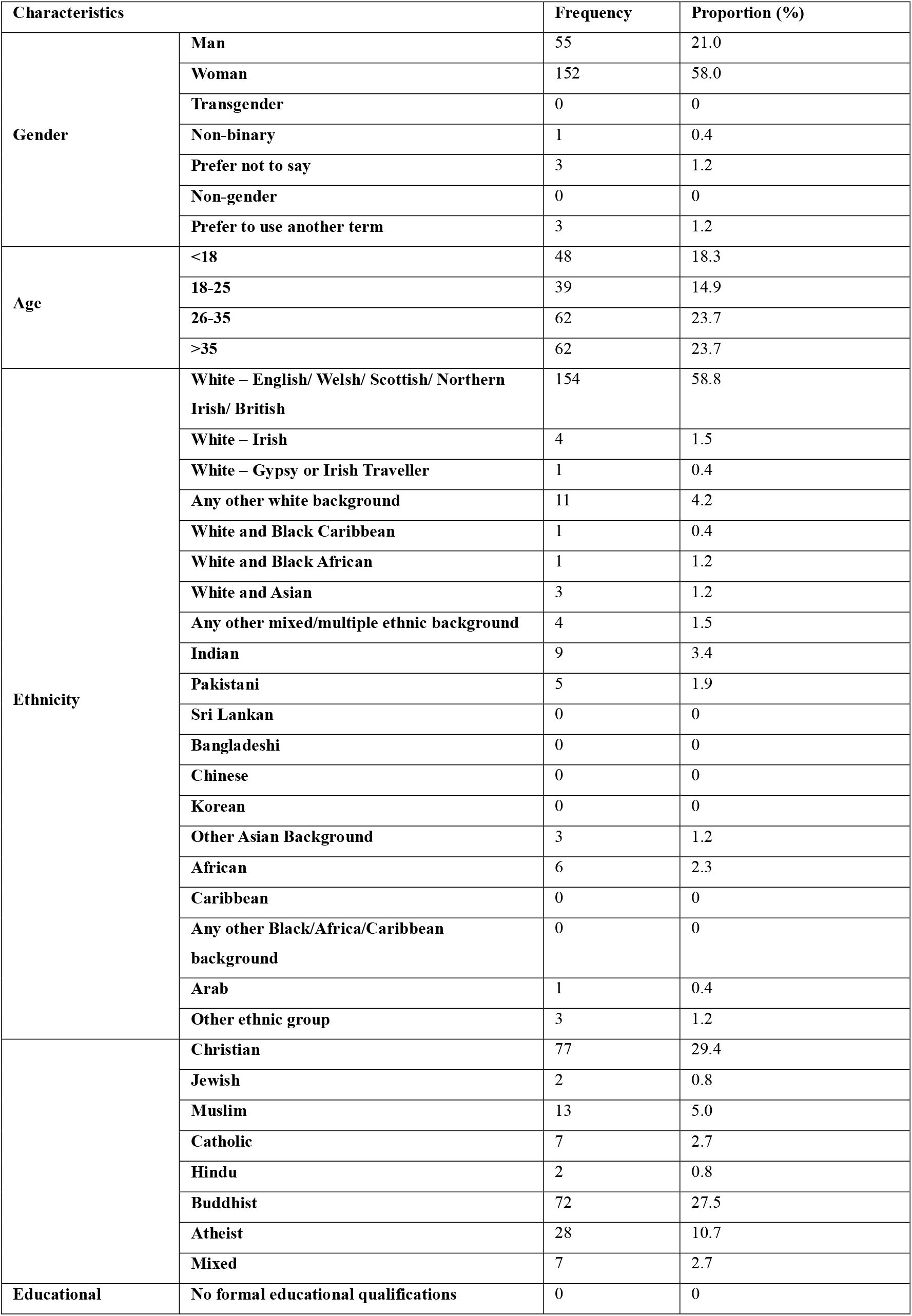
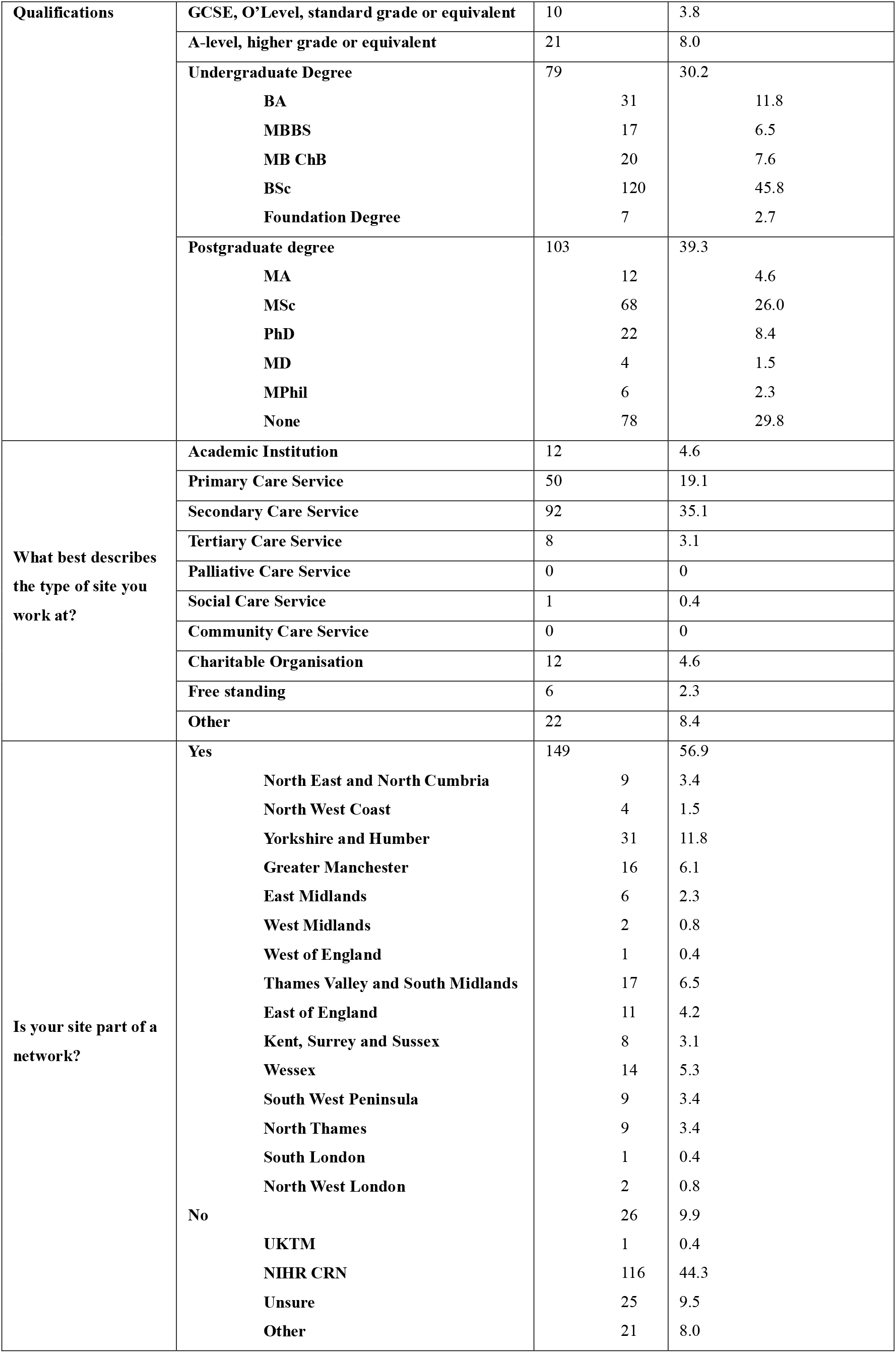
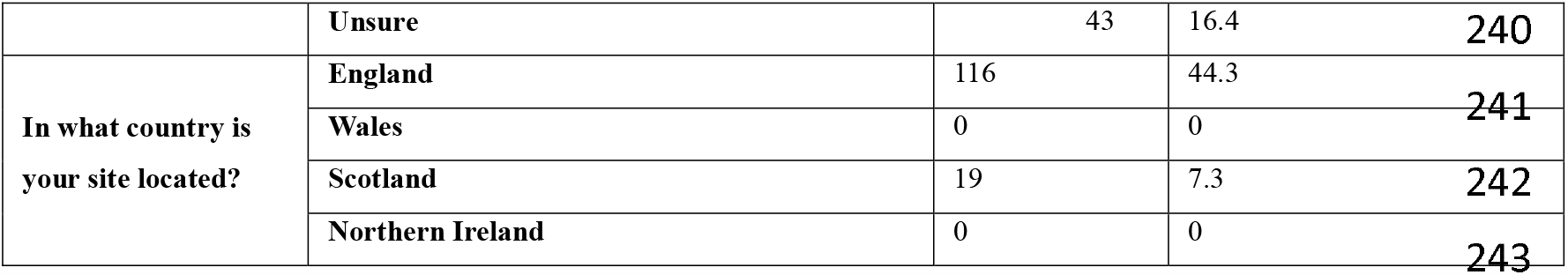
UK Demographics.

**Table 1b.** UK Demographics.

|  |  | US Site 1 |  | US Site 2 |  | Total US |  |
| --- | --- | --- | --- | --- | --- | --- | --- |
|  |  | Frequency | Proportion (%) | Frequency | Proportion (%) | Frequency | Proportion (%) |
| What best describes the type of site you work at? | Academic | 48 | 16.6 | 1 | 0.4 | 49 | 8.6 |
|  | Freestanding Research Center | 132 | 45.5 | 166 | 59.7 | 298 | 52.5 |
|  | Hospital | 37 | 12.8 | 5 | 1.8 | 42 | 7.4 |
|  | Physician Practice/Clinic | 73 | 25.2 | 106 | 38.2 | 179 | 31.5 |
|  | Total | 290 | 100 | 278 | 100 | 568 | 100 |
| Is the site part of a network? | Yes | 129 | 44.2 | 113 | 40.9 | 242 | 42.6 |
|  | No | 135 | 46.6 | 163 | 59.1 | 298 | 52.5 |
|  | Unsure | 28 | 9.6 | - | - | 28 | 4.9 |
|  | Total | 292 | 100 | 276 | 100 | 568 | 100 |
| What best describes your role? | Assistant | 5 | 1.7 | - | - | 5 | 0.9 |
|  | Director/ Vice President | 51 | 17.5 | - | - | 51 | 9 |
|  | Investigator | 36 | 12.3 | 66 | 23.7 | 102 | 17.9 |
|  | Owner/President | 24 | 8.2 | - | - | 24 | 4.2 |
|  | Regulatory/Compliance | 8 | 2.7 | - | - | 8 | 1.4 |
|  | Research Assistant | - | - | 3 | 1.1 | 3 | 0.5 |
|  | Site Manager | 71 | 24.3 | 97 | 34.9 | 168 | 29.5 |
|  | Study Co-ordinator | 50 | 17.1 | 28 | 10.1 | 78 | 13.7 |
|  | Other | 47 | 16.1 | 84 | 30.2 | 131 | 23 |
|  | Total | 292 | 100 | 278 | 100 | 570 | 100 |
| How many studies is your site currently conducting? | Less than 10 | 116 | 39.7 | 147 | 52.9 | 263 | 46.1 |
|  | 10-20 | 69 | 23.6 | 85 | 30.6 | 154 | 27.0 |
|  | 20-30 | 34 | 11.6 | 30 | 10.8 | 64 | 11.2 |
|  | 30+ | 73 | 25.0 | 16 | 5.8 | 89 | 15.6 |
|  | Total | 292 | 100 | 278 | 100 | 570 | 100 |

Data from all participants was used for the descriptive analysis presented in Table 3. Since some participants did not complete all items across all sections, care was taken during the categorical and comparative analysis to use data for only those participants who had effectively completed most items across all sections to make comparisons meaningful. For comparative analysis, this resulted in an effective response from 703 participants: US Cohort 1 (N=229), US Cohort 2 (N=293) and UK Cohort (N=181). Table 2 presents the results of the reliability analysis. This shows the Cronbach’s alpha and the average interitem correlations for all participants and cohorts from both US and UK indicate that the DSAT and items within its three sections have high reliability.

**Table 2.** Reliability Analysis.

| DSAT Element | UK Cohort |  | US Cohorts |  | All |  |
| --- | --- | --- | --- | --- | --- | --- |
|  | *α* | Average ICC | *α* | Average ICC | *α* | Average ICC |
| Site Overview (10 items) | 0.89 | 0.46 | 0.90 | 0.48 | 0.90 | 0.48 |
| Site Recruitment and Outreach (9 items) | 0.86 | 0.40 | 0.91 | 0.54 | 0.91 | 0.52 |
| Patient Focused Services (6 items) | 0.61 | 0.23 | 0.70 | 0.31 | 0.72 | 0.33 |
| DSAT (25 item) | 0.90 | 0.28 | 0.94 | 0.41 | 0.94 | 0.40 |

**Table 3.** DSAT Scores of US and UK Study Participants.

| DSAT | N | Mean | SD |
| --- | --- | --- | --- |
| Section I. Site Overview | **852** | **40.97** | **21.35** |
| Our site knows the demographic make-up of our community | 703 | 5.24 | 1.20 |
| Site knows the actual patient demographics of the clinical trial participants | 703 | 5.19 | 1.34 |
| Investigator, Study Coordinator, and/or support staff are culturally representative of the patient population | 702 | 4.81 | 1.38 |
| Site has staff that can utilize commonly used words in the community for medical discussion | 702 | 5.17 | 1.35 |
| Site staff knows diversity goals | 702 | 5.06 | 1.47 |
| Site tracks progress toward established diversity goals and knows what marketing or outreach strategies works to make them successful | 701 | 4.35 | 1.76 |
| Site can provide diversity metrics upon request to sponsor at site evaluation | 699 | 4.81 | 1.66 |
| Site has mechanisms (formal or informal) to make staff culturally aware and sensitive to the patient population | 699 | 4.95 | 1.49 |
| Site management team supports the recruitment of diverse patients | 701 | 5.44 | 1.27 |
| Site has a process to identify and address barriers | 701 | 4.76 | 1.59 |
| Section II. Site Recruitment and Outreach | **851** | **33.71** | **19.93** |
| Site has an established tailored strategy to approach targeted populations for clinical trials | 682 | 4.83 | 1.56 |
| When needed, site partners with sponsor to create study-specific and target patient population specific recruitment plans | 682 | 4.62 | 1.73 |
| Site creates target patient population specific retention plans | 681 | 4.40 | 1.87 |
| When needed, site requests appropriately tailored patient materials from sponsor | 681 | 5.03 | 1.57 |
| Site routinely requests materials (e.g., ICF, patient recruitment materials, patient outcomes assessments, E-diaries) in languages predominantly used by members of their patient population | 680 | 4.85 | 1.73 |
| When needed, site conducts outreach to minority-based organizations to establish a network of referrals (e.g., churches, community centers, food banks, medical community, patient advocacy and support groups, etc.) | 680 | 4.16 | 1.90 |
| When needed, site utilizes appropriate media outlets (radio, TV, social media) specific to the targeted population | 680 | 4.40 | 1.86 |
| Site uses recruitment materials that have worked in the past to decide upon future strategy for recruiting diverse patients | 679 | 4.69 | 1.73 |
| Site has a mechanism to notify patients for eligibility in clinical trials | 680 | 5.18 | 1.45 |
| Section III. Patient Focused Services | **849** | **20.63** | **12.59** |
| Informed Consent form is available, pre-translated and pre-IRB approved in relevant languages | 675 | 4.73 | 1.79 |
| If allowed by state/local regulations, site accommodates working patients by offering hours of operation outside of normal business hours (i.e., early arrivals and/or after hours appointments) and weekend visits | 673 | 4.70 | 1.62 |
| Site offers phone prompt for most frequently used languages (i.e., press 1 for Spanish) | 674 | 3.50 | 2.56 |
| Site provides transportation services or requests reimbursement from sponsor as needed | 674 | 4.90 | 1.59 |
| When needed, site has provisions for providing a place to stay for patients and their family members including children | 674 | 3.33 | 2.31 |
| Stipends are offered and/or distributed in a timely manner and method easy for patient use | 673 | 4.88 | 1.75 |

Table 3 and Figure 1 provide a descriptive insight into the overall DSAT score and Section scores when all participants were included in the analysis. Based on the mean scores, it can be inferred that on an average, members report that their sites <u>often</u> engaged in activities associated with site overview and site recruitment and outreach best practices, but relatively less frequent (<u>sometimes)</u> in activities associated with patient focused services. Unsurprisingly, the categorical distribution showcased in Figure 1 indicates that 82% members reported always or nearly always experiencing site overview best practices, 70% members reported always or nearly always experiencing site recruitment and outreach best practices but only 59% members reported always or nearly always experiencing patient focused services best practices. Items in the Patient Focused Services section such as “When needed, site has provisions for providing a place to stay for patients and their family members including children” and “Site offers phone prompt for most frequently used languages (i.e., press 1 for Spanish)” had the lowest mean scores across all three sections.

**Figure 1.**
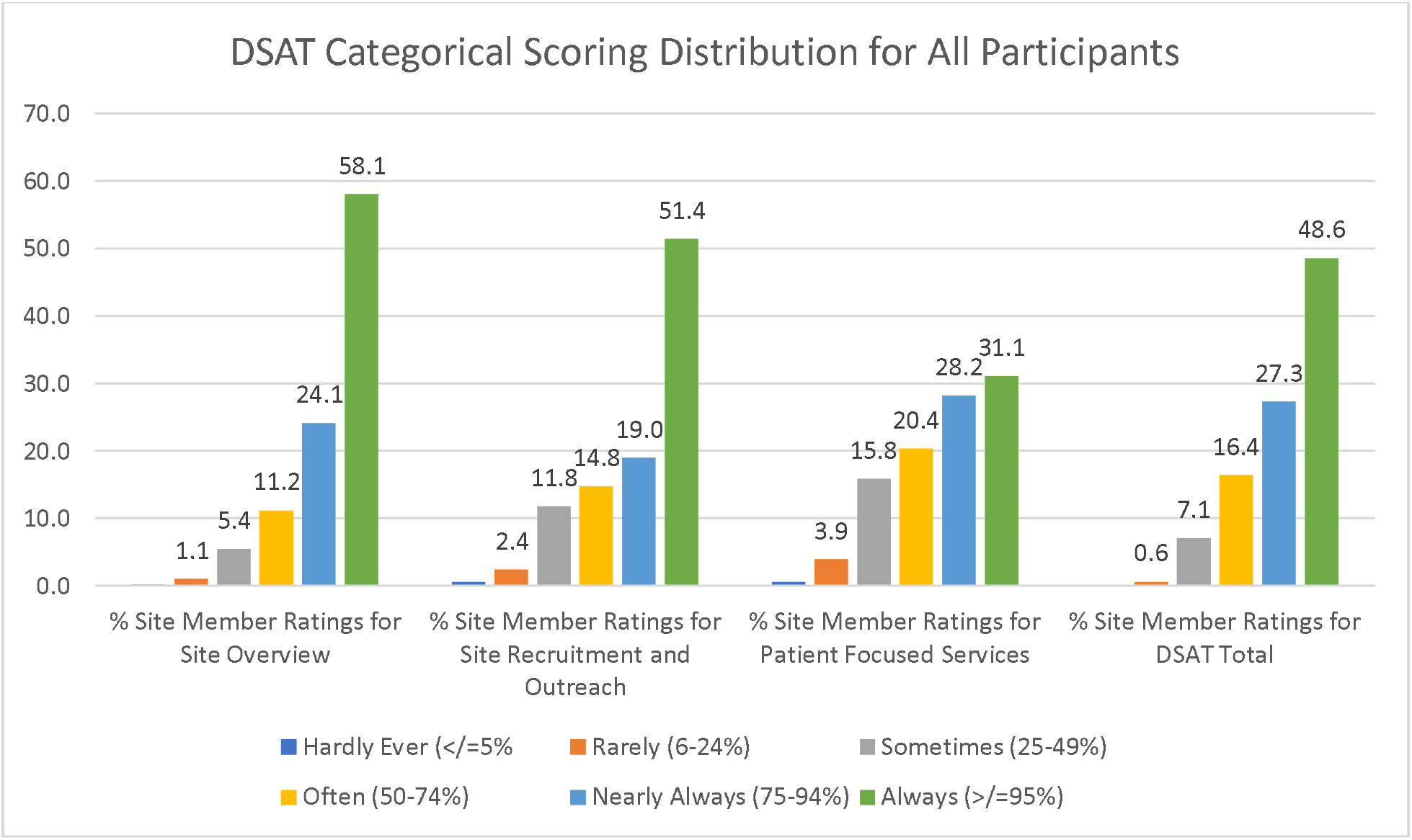
DSAT Categorical Scoring Distribution of All Participants.

In Table 4, a comparison of the mean scores for the combined US cohorts with the UK cohorts and the p-values associated with each item on the DSAT is provided. As can be seen, US sites average scores on all 3 sections were closer to 5 (nearly always) whereas UK sites average scores were closer to 4 (often). The mean scores of US sites were significantly higher than UK sites on 23 items and only 2 items (item 8 in Site Overview and item 3 Patient Focused Services) showed no significant difference in means between US and UK cohorts. Three items from the Patient Focused Services section (item 1, item 5 and item 5) and two items from the Site Recruitment and Outreach (item 3 and 5) show the highest difference between US and UK sites. Figures 2a and 2b provide a graphical illustration of the categorical distribution of DSAT sections and total DSAT scores. It is interesting to note that there is at least a 20% difference in percentage of members who indicated nearly always/always in all 3 sections and the total DSAT, with the highest difference between the US and UK cohorts being on the Patient Focused Services section. (88% US vs 65% UK on Site Overview; 78% US vs 48% UK on Site Recruitment and Outreach; 71% US vs 26% for UK on Patient Focused Services; 83% US vs 54% UK on overall DSAT).

**Table 4.**
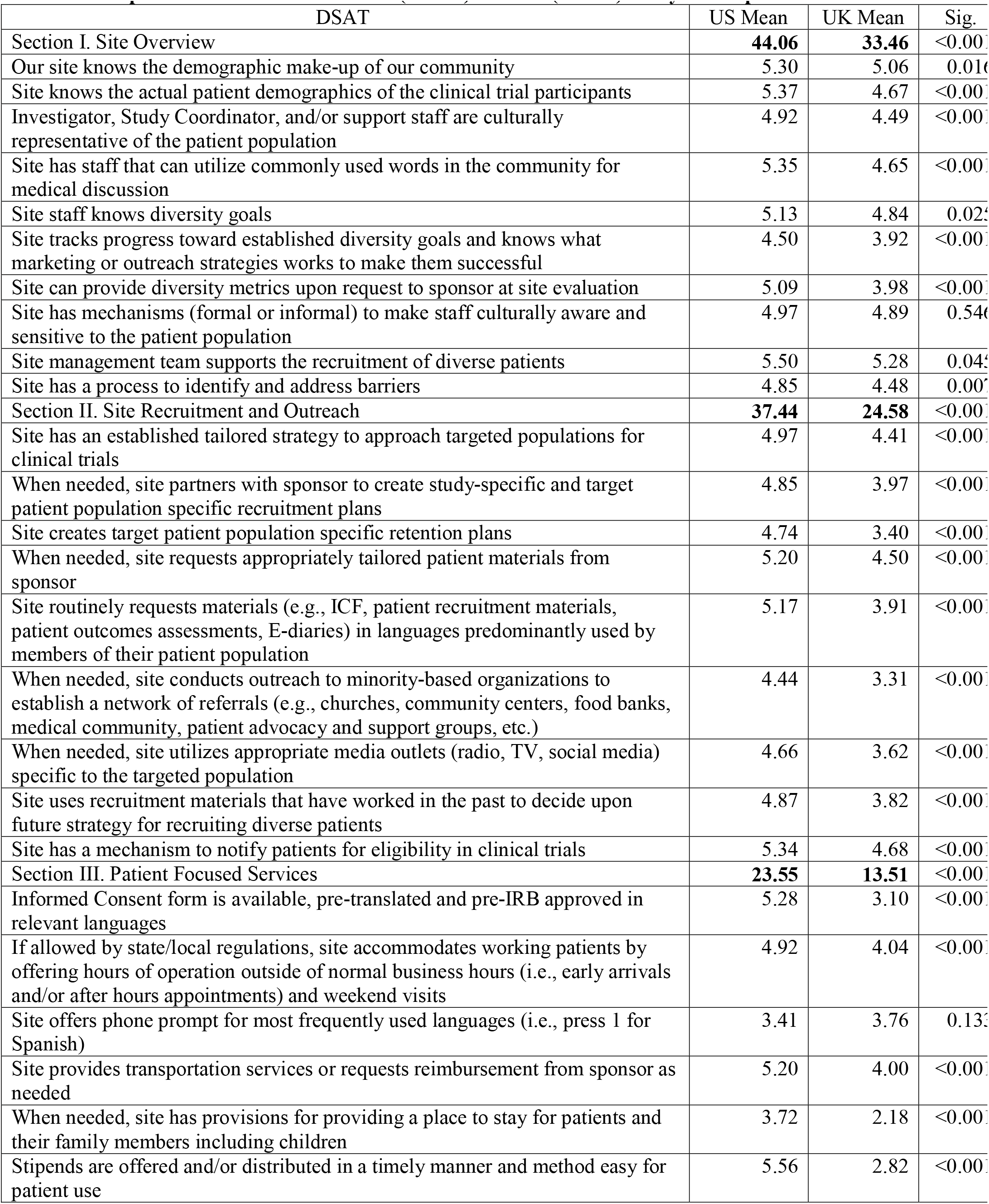
Comparison of DSAT Scores of US (N=522) and UK (N=181) Study Participants.

| DSAT | US Mean | UK Mean | Sig. |
| --- | --- | --- | --- |
| Section I. Site Overview | **44.06** | **33.46** | <0.001 |
| Our site knows the demographic make-up of our community | 5.30 | 5.06 | 0.016 |
| Site knows the actual patient demographics of the clinical trial participants | 5.37 | 4.67 | <0.001 |
| Investigator, Study Coordinator, and/or support staff are culturally representative of the patient population | 4.92 | 4.49 | <0.001 |
| Site has staff that can utilize commonly used words in the community for medical discussion | 5.35 | 4.65 | <0.001 |
| Site staff knows diversity goals | 5.13 | 4.84 | 0.025 |
| Site tracks progress toward established diversity goals and knows what marketing or outreach strategies works to make them successful | 4.50 | 3.92 | <0.001 |
| Site can provide diversity metrics upon request to sponsor at site evaluation | 5.09 | 3.98 | <0.001 |
| Site has mechanisms (formal or informal) to make staff culturally aware and sensitive to the patient population | 4.97 | 4.89 | 0.546 |
| Site management team supports the recruitment of diverse patients | 5.50 | 5.28 | 0.045 |
| Site has a process to identify and address barriers | 4.85 | 4.48 | 0.007 |
| Section II. Site Recruitment and Outreach | **37.44** | **24.58** | <0.001 |
| Site has an established tailored strategy to approach targeted populations for clinical trials | 4.97 | 4.41 | <0.001 |
| When needed, site partners with sponsor to create study-specific and target patient population specific recruitment plans | 4.85 | 3.97 | <0.001 |
| Site creates target patient population specific retention plans | 4.74 | 3.40 | <0.001 |
| When needed, site requests appropriately tailored patient materials from sponsor | 5.20 | 4.50 | <0.001 |
| Site routinely requests materials (e.g., ICF, patient recruitment materials, patient outcomes assessments, E-diaries) in languages predominantly used by members of their patient population | 5.17 | 3.91 | <0.001 |
| When needed, site conducts outreach to minority-based organizations to establish a network of referrals (e.g., churches, community centers, food banks, medical community, patient advocacy and support groups, etc.) | 4.44 | 3.31 | <0.001 |
| When needed, site utilizes appropriate media outlets (radio, TV, social media) specific to the targeted population | 4.66 | 3.62 | <0.001 |
| Site uses recruitment materials that have worked in the past to decide upon future strategy for recruiting diverse patients | 4.87 | 3.82 | <0.001 |
| Site has a mechanism to notify patients for eligibility in clinical trials | 5.34 | 4.68 | <0.001 |
| Section III. Patient Focused Services | **23.55** | **13.51** | <0.001 |
| Informed Consent form is available, pre-translated and pre-IRB approved in relevant languages | 5.28 | 3.10 | <0.001 |
| If allowed by state/local regulations, site accommodates working patients by offering hours of operation outside of normal business hours (i.e., early arrivals and/or after hours appointments) and weekend visits | 4.92 | 4.04 | <0.001 |
| Site offers phone prompt for most frequently used languages (i.e., press 1 for Spanish) | 3.41 | 3.76 | 0.135 |
| Site provides transportation services or requests reimbursement from sponsor as needed | 5.20 | 4.00 | <0.001 |
| When needed, site has provisions for providing a place to stay for patients and their family members including children | 3.72 | 2.18 | <0.001 |
| Stipends are offered and/or distributed in a timely manner and method easy for patient use | 5.56 | 2.82 | <0.001 |

**Figure 2a.**
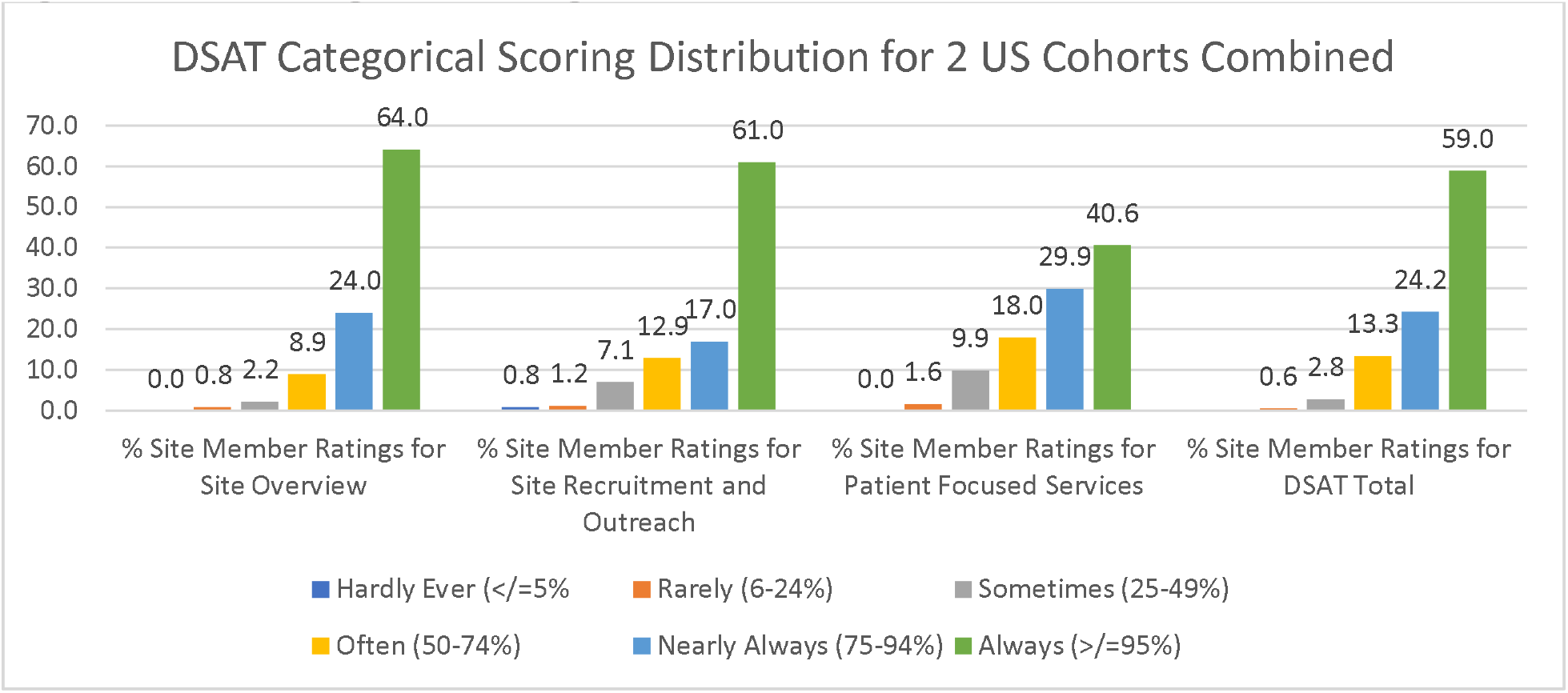
DSAT Categorical Scoring Distribution of US Cohorts.

**Figure 2b.**
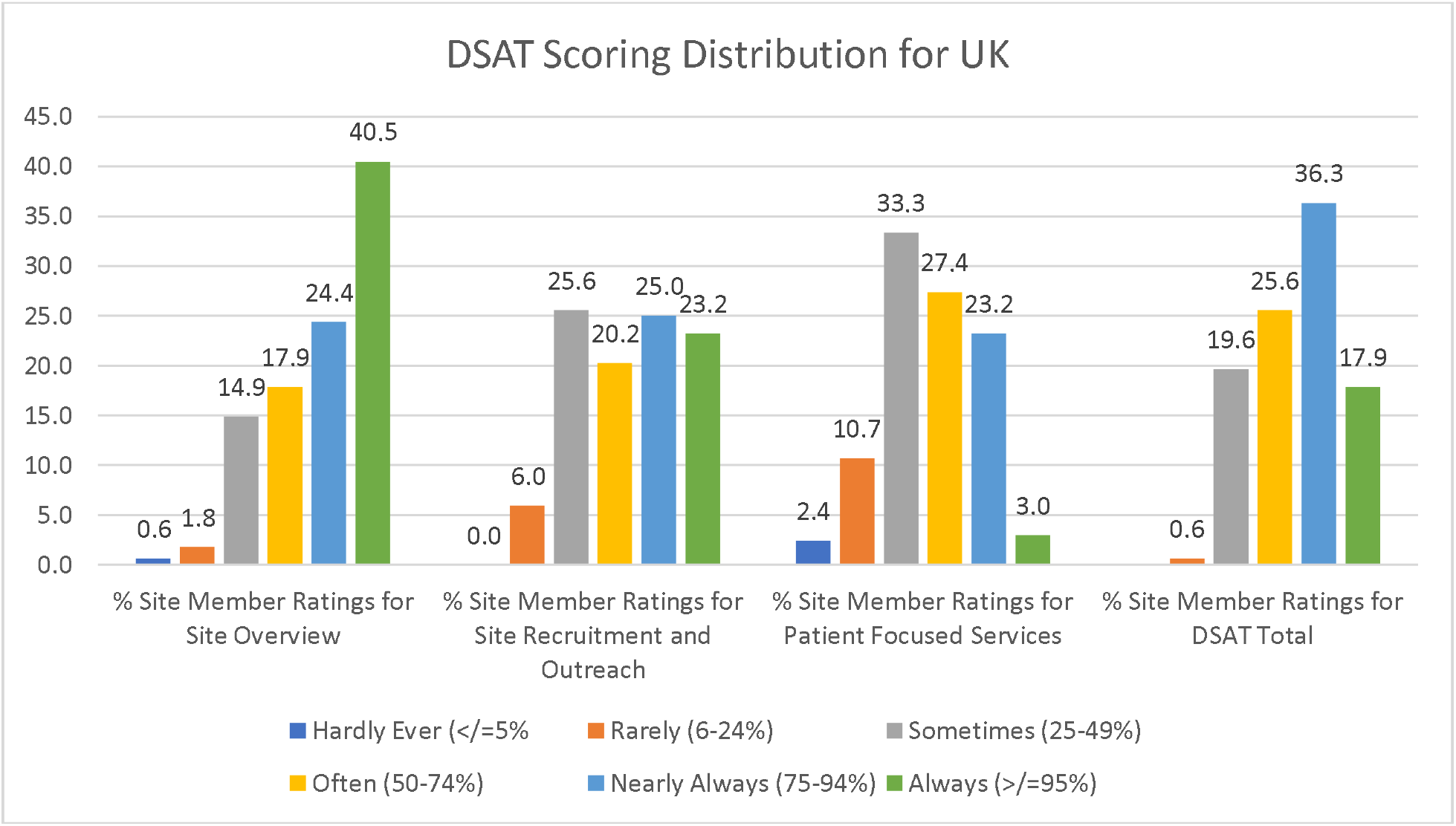
DSAT Categorical Scoring Distribution of US Cohorts.

To further delve into the differences between the US and UK cohort responses, mean scores of the UK cohort was compared with segregated data from each of the US cohorts (US cohort 1 and US cohort 2). Table 5, Figure 3a, Figure 3b and the previously presented Figure 2b provide these comparisons. When mean scores of the three cohorts are examined, it becomes clear that the mean scores for US cohort 2 are significantly higher than those compared to mean scores for US cohort 1 and the UK cohort. It is also interesting to note that mean scores for the US 1 cohort were lower than the UK cohort for only 8 out of 25 items (7 items in the Site Overview section and 1 item from the Patient Focused Services section). Overall, the highest difference between US cohort 2 and the UK cohort means were on 3 items in the Patient Focused Services section (items 1,5, and 6) and two items in the Site Recruitment and Outreach section items (items 3 and 6). Overall highest difference between US cohort 1 and US cohort 2 were on 1 item in the Site Overview section (item 6) and 3 items in the Site Recruitment and Outreach section (items 3, 6, and 7). The graphics from Figure 3a, 3b and 2 b clearly indicate the percentage of members from US cohort 2 who indicated nearly always/always in all 3 sections and the total DSAT was significantly higher than US cohort 1 and the UK cohort, wherein the UK cohort had the lowest percentage (80% US 1, 94% US 2 vs 65% UK on Site Overview; 63% US 1, 89% US 2 vs 48% UK on Site Recruitment and Outreach; 57% US 1, 80% US 2 vs 26% for UK on Patient Focused Services; 69% US 1, 94% US 2 vs 54% UK on total DSAT).

**Table 5.** Comparison of DSAT Scores By Data Source (US 1 (N=229), US 2 (N=293) and UK (N=181)

| DSAT | US1<br>Mean | US 2<br>Mean | UK<br>Mean | Sig |
| --- | --- | --- | --- | --- |
| Section I. Site Overview | **36.26** | **51.51** | **33.46** | <0.001 |
| Our site knows the demographic make-up of our community | 5.01 | 5.53 | 5.06 | <0.001 |
| Site knows the actual patient demographics of the clinical trial participants | 5.01 | 5.66 | 4.67 | <0.001 |
| Investigator, Study Coordinator, and/or support staff are culturally representative of the patient population | 4.59 | 5.18 | 4.49 | <0.001 |
| Site has staff that can utilize commonly used words in the community for medical discussion | 5.04 | 5.58 | 4.65 | <0.001 |
| Site staff knows diversity goals | 4.64 | 5.51 | 4.84 | <0.001 |
| Site tracks progress toward established diversity goals and knows what marketing or outreach strategies works to make them successful | 3.87 | 4.99 | 3.92 | <0.001 |
| Site can provide diversity metrics upon request to sponsor at site evaluation | 4.56 | 5.51 | 3.98 | <0.001 |
| Site has mechanisms (formal or informal) to make staff culturally aware and sensitive to the patient population | 4.44 | 5.38 | 4.89 | <0.001 |
| Site management team supports the recruitment of diverse patients | 5.24 | 5.70 | 5.28 | <0.001 |
| Site has a process to identify and address barriers | 4.31 | 5.28 | 4.48 | <0.001 |
| Section II. Site Recruitment and Outreach | **28.82** | **45.67** | **24.58** | <0.001 |
| Site has an established tailored strategy to approach targeted populations for clinical trials | 4.44 | 5.36 | 4.41 | <0.001 |
| When needed, site partners with sponsor to create study-specific and target patient population specific recruitment plans | 4.33 | 5.23 | 3.97 | <0.001 |
| Site creates target patient population specific retention plans | 3.99 | 5.30 | 3.40 | <0.001 |
| When needed, site requests appropriately tailored patient materials from sponsor | 4.73 | 5.55 | 4.50 | <0.001 |
| Site routinely requests materials (e.g., ICF, patient recruitment materials, patient outcomes assessments, E-diaries) in languages predominantly used by members of their patient population | 4.78 | 5.46 | 3.91 | <0.001 |
| When needed, site conducts outreach to minority-based organizations to establish a network of referrals (e.g., churches, community centers, food banks, medical community, patient advocacy and support groups, etc.) | 3.72 | 4.98 | 3.31 | <0.001 |
| When needed, site utilizes appropriate media outlets (radio, TV, social media) specific to the targeted population | 3.91 | 5.21 | 3.62 | <0.001 |
| Site uses recruitment materials that have worked in the past to decide upon future strategy for recruiting diverse patients | 4.41 | 5.39 | 3.82 | <0.001 |
| Site has a mechanism to notify patients for eligibility in clinical trials | 4.88 | 5.69 | 4.68 | <0.001 |
| Section III. Patient Focused Services | **18.54** | **28.33** | **13.51** | <0.001 |
| Informed Consent form is available, pre-translated and pre-IRB approved in relevant languages | 4.81 | 5.62 | 3.10 | <0.001 |
| If allowed by state/local regulations, site accommodates working patients by offering hours of operation outside of normal business hours (i.e., early arrivals and/or after hours appointments) and weekend visits | 4.59 | 5.19 | 4.04 | <0.001 |
| Site offers phone prompt for most frequently used languages (i.e., press 1 for Spanish) | 2.90 | 3.79 | 3.76 | <0.001 |
| Site provides transportation services or requests reimbursement from sponsor as needed | 4.81 | 5.48 | 4.00 | <0.001 |
| When needed, site has provisions for providing a place to stay for patients and their family members including children | 3.18 | 4.12 | 2.18 | <0.001 |
| Stipends are offered and/or distributed in a timely manner and method easy for patient use | 5.21 | 5.82 | 2.82 | <0.001 |

**Figure 3a.**
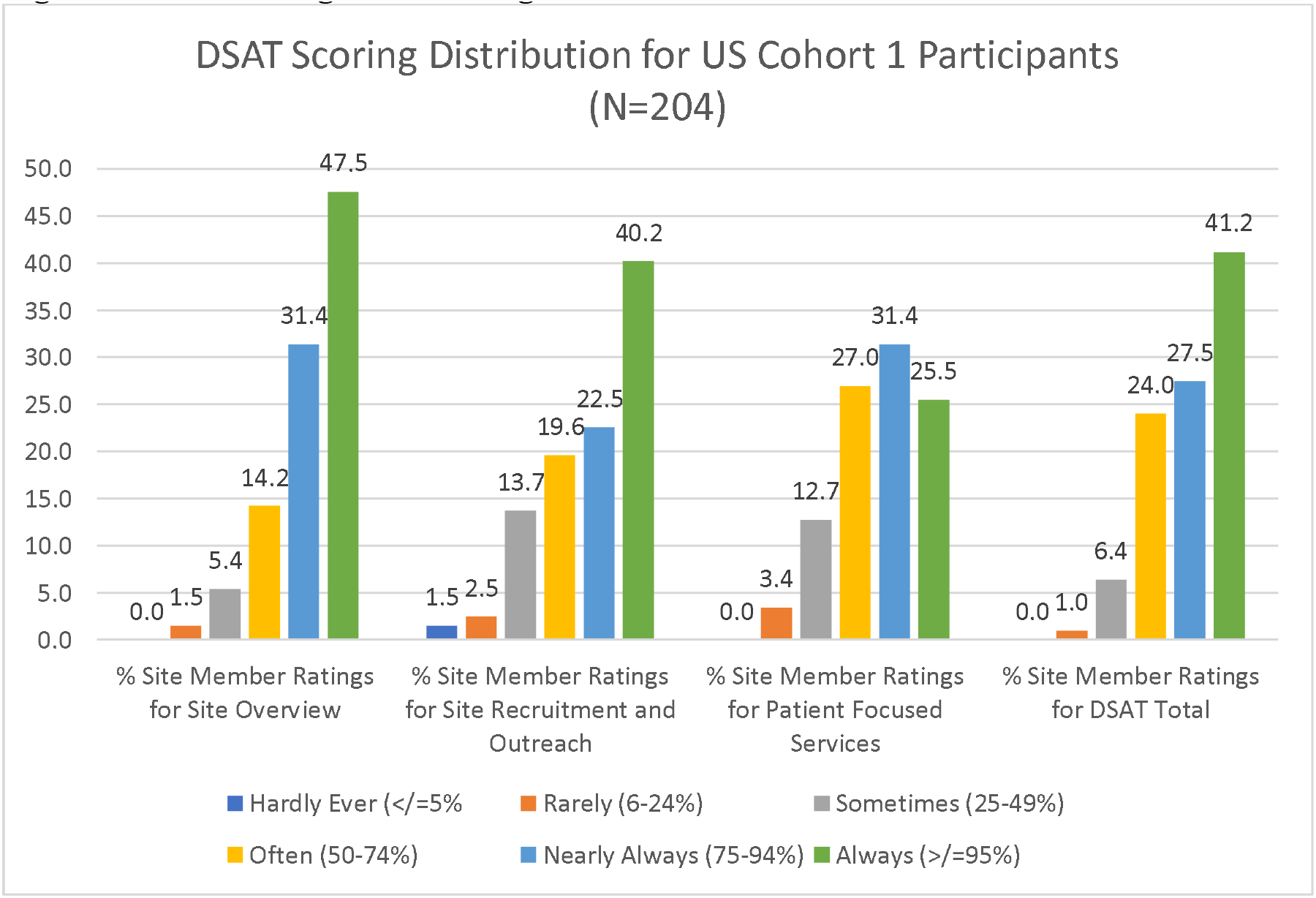
DSAT Categorical Scoring Distribution of US Cohort 1.

**Figure 3b.**
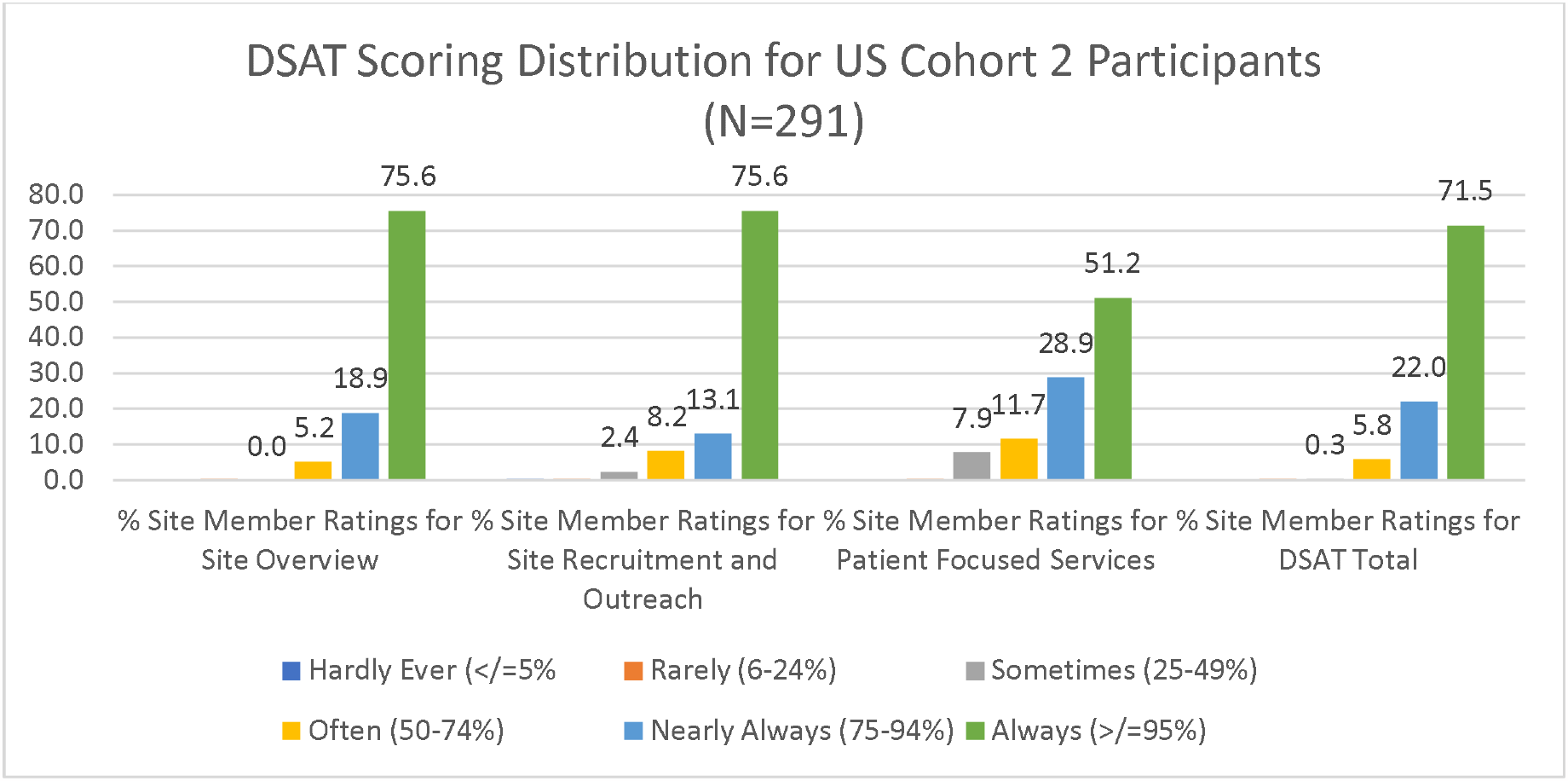
DSAT Categorical Scoring Distribution of US Cohort 1.

## Discussion

Globally, there is a movement underway to improve diversity of populations participating in clinical trials. Governmental and regulatory agencies, non-profits and private enterprises involved in discovery, development and approval of pharmaceutical products are clearly expending efforts towards greater representation of diverse ethnic populations and groups that are usually underrepresented in clinical trials. In such an environment, the availability of a self-assessment instrument such as the DSAT has been identified as providing an opportunity to utilize a continuous quality improvement and collaborative approach to diversity recruitment, outreach, and management in clinical trials. As prior published studies using the DSAT primarily showcased perspectives of site members from the United States, there was a need to examine the utility of DSAT in other countries and provide comparative data on the status of practices related to diversity recruitment, outreach, and management in clinical trials. This study is a step towards addressing the gap.

There are several noteworthy findings from this study which would be an excellent utility to the clinical trials industry and those involved in partnering with it. First, the reliability of the DSAT was consistently high across all 3 cohorts that utilized it. This finding is important because it showcases the fact that the instrument can be used in non-US contexts for the self-assessment of best practices involved in clinical trials industry as it pertains to diversity recruitment, outreach and management. Second, the descriptive findings of the study clearly indicate that practices associated with patient focused services such as phone prompts for language services, providing access to a place for patient’s families and children to stay are areas that can be improved upon. The findings also indicate that outreach and recruitment activities are sites are still not optimally conducting outreach to minority-based organisations to establish a network of referrals and using appropriate media outlets as well as creating target population specific retention plans. These findings indicate that clinical trial sponsors and sites may need some assistance and training in these areas of best practices. It was both interesting and heartening to note that the highest mean score was associated with the item “Site management team supports the recruitment of diverse patients”, which indicates that a culture of diversity recruitment, outreach and management is prevalent in the industry. The third and an important finding from this study was that the DSAT scores (all three sections and total) for the US cohort was significantly higher than the UK cohort. While not investigated in this study, a potential reason for this finding could be related to the history of how the FDA, the industry, and its stakeholders such as SCRS have engaged in activities that may have spurred the early adoption of best practices in improving diversity in clinical trials. Alternatively, another reason could be that the norms associated with clinical trials recruitment, outreach, and management as measured in the DSAT were shaped in the US and are now slowly being adopted in the UK. Different stakeholders from US and UK might want to examine the findings carefully and discuss its implications for their own country and sites within it.

Finally, it was interesting to note the difference in the DSAT scores between the two US cohorts. Future case studies may want to explore and investigate how best practices in diversity are being integrated in different countries and in different states within a regionally large and diverse country such as the US. There are several limitations that should be considered when taking these findings into consideration. This study was based on a secondary analysis of data from cohorts of members that were involved in this study. These cohorts participated during different time frame and had access to the DSAT via different mechanisms depending on how they were invited or recruited. Primary research studies using study designs that will provide for simultaneous access and participation are needed.

## Conclusion

This study provides an important insight to the utility of DSAT for cross-country comparisons. It found that overall, the best practices associated with site management, site recruitment and outreach of diverse populations are nearly always utilized whereas practices associated with patient focused services are sometimes or utilized often. It also found that best practices identified by DSAT are utilized at a higher rate in clinical trials sites in the United States as compared to sites in the United Kingdom.

## Data Availability

All data produced in the present study are available upon reasonable request to the authors

## Notes

### Competing Interest Statement

Dr Diane Foster developed the DSAT tool. PP has received research grant from Novo Nordisk, and other, educational from Queen Mary University of London, other from John Wiley & Sons, other from Otsuka, outside the submitted work. All other authors report no relevant conflicts of interest for this article.

### Author Declarations

HRA and Health and Care Research Wales (HCRW) Approval.

Application does not require HRA/HCRW and REC Approval in order to proceed at participating NHS organisations in England or Wales.

